# Deep clinical phenotyping of Alzheimer’s Disease Patients Leveraging Electronic Medical Records Data Identifies Sex-Specific Clinical Associations

**DOI:** 10.1101/2021.03.19.21253659

**Authors:** Alice Tang, Tomiko Oskotsky, William Mantyh, Caroline Warly Solsberg, Billy Zeng, Zicheng Hu, Boris Oskotsky, Dena Dubal, Marina Sirota

**Affiliations:** Bakar Computational Health Sciences Institute, UCSF, San Francisco, CA, USA; Graduate Program in Bioengineering, UCSF, San Francisco, CA, USA; School of Medicine, UCSF, San Francisco, CA, USA; Department of Pediatrics, UCSF, San Francisco, CA, USA; Department of Neurology, University of Minnesota School of Medicine, Delaware, MN, USA; Pharmaceutical Sciences and Pharmacogenomics, UCSF, San Francisco, CA, USA; Department of Neurology and Weill Institute for Neurosciences, University of California, San Francisco, San Francisco, CA 94158, USA

**Author notes:** Correspondence should go to or.

## Abstract

Alzheimer’s Disease (AD) is a devastating disorder that is still not fully understood. Sex modifies AD vulnerability, but the reasons for this are largely unknown. There has been efforts to understand select comorbidities, covariates, and biomarkers of AD, with and without sex stratification - but there has not yet been an integrative, big data approach to identify clinical and sex specific associations with AD in an unbiased manner. Electronic Medical Records (EMR) contain extensive information on patients, including diagnoses, medications, and lab test results, providing a unique opportunity to apply phenotyping approaches to derive insights into AD clinical associations. Here, we utilize EMRs to perform deep clinical phenotyping and network analysis of AD patients to provide insight into its clinical characteristics and sex-specific clinical associations. We performed embeddings and network representation of patient diagnoses to visualize patient heterogeneity and comorbidity interactions and observe greater connectivity of diagnosis among AD patients compared to controls. We performed enrichment analysis between cases and controls and identified multiple known and new diagnostic and medication associations, such as positive associations with AD and hypertension, hyperlipidemia, anemia, and urinary tract infection - and negative associations with neoplasms and opioids. Furthermore, we performed sex-specific enrichment analyses to identify novel sex-specific associations with AD, such as osteoporosis, depression, cardiovascular risk factors, and musculoskeletal disorders diagnosed in female AD patients and neurological, behavioral, and sensory disorders enriched in male AD patients. We also analyzed lab test results, resulting in clusters of patient phenotype groups, and we observed greater calcium and lower alanine aminotransferase (ALT) in AD, as well as abnormal hemostasis labs in female AD. With this method of phenotyping, we can represent AD complexity, and identify clinical factors that can be followed-up for further temporal and predictive analysis or integrate with molecular data to aid in diagnosis and generate hypotheses about disease mechanisms. Furthermore, the negative associations can help identify factors that may decrease likelihood of AD and help motivate future drug repurposing or therapeutic approaches.

## Introduction

Alzheimer’s disease (AD) is the most common type of dementia, making up 60-80% of cases, with devastating impact on patients’ lives and projections of being an increasing burden for the future^1^. AD is characterized by brain atrophy and accumulation of beta-amyloid plaques and tau tangles seen on brain pathology after death. The disease erodes memory and cognitive functions, causing interference with daily activities and contributing to great emotional, social, and economic burden on patients and their families, yet the disease remains incurable and challenging to understand and diagnose. One reason AD is difficult to study is because it is a complex, heterogenous, and multifactorial disease that takes many years to manifest^2^. This complexity, along with the slow insidious progression of AD, makes it difficult to fully characterize disease phenotypes and associations.

Sex is one factor that has been shown to be important in AD, with higher prevalence in women afflicted by the disease at a 2:1 ratio^1^. Recent findings show that sex contributes to differing vulnerabilities or resilience to AD, as men progress to death quicker^3, 4^ while women show higher cognitive resilience despite increased tau pathology^3, 5, 6^. How sex contributes to these differences in prevalence and vulnerability is a question of fervent interest among researchers in the AD field^7^. Recent studies in mice demonstrate that a second X chromosome may contribute to AD resilience^4^. Further sex-specific studies in AD also show sex modification of AD risk^8^, progression^9^, and molecular phenotype^9–12^. As such, sex is an important factor to consider in studying and phenotyping AD.

While many efforts have evaluated the association of individual risk factors with AD, unbiased approaches to these associations are limited. Prior work, largely hypothesis-driven, focused on select comorbidities associated with AD, such as hypertension^13^, vascular disorders^14^, diabetes^15^, obesity^16^, and others^17–19^. However, how sex modultes AD complexity and heterogeneity has still not been fully explored. Prior big data approaches to AD have examined genotype-phenotype associations^20, 21^ and molecular analyses^12, 22, 23^ to characterize AD disease and sex differences^10, 11^. Other work on phenotyping AD patients using clinical data has examined neuroimaging^24^, neuropsychiatric phenotype^25^, chart reviews^26^, and billing records independently. Thus, an unbiased comprehensive approach to phenotype AD and identify sex associations using full clinical records is needed.

With an abundance of electronic medical record (EMR) use over the past decade, there is extensive underutilized clinical data on patients covering comorbidities, medications, and lab values. This type of dataset provides a great opportunity to deeply investigate diseases and identify associations to facilitate understanding disease prevention and progression. Recently, EMR has been utilized for other diseases for creating comorbidity networks^27^, identifying disease subtypes^28^ and predicting disease outcomes^29, 30^ highlighting the potential of utilizing EMR data to extract insight and utility for complex and heterogeneous diseases^31^, but a big data integrative analysis with EMR data has not yet been applied to the AD phenotype.

Here, we take an integrative approach through deep clinical phenotyping and network analysis to provide insight into AD clinical characteristics with a focus on sex differences. For the first time, integrative phenotyping and association analysis is used to identify, in an unbiased manner, unique clinical features associated with AD itself - and reveals previously unknown sex-specific associations in the context of diagnosed, medications and lab test results.

## Results

### Cohort Selection and Visualization

From the UCSF EMR database (∼5 million patients), we identified 8,804 AD patients (5,558 females and 3,220 males) and 17,608 propensity score-matched control patients (11,117 females and 6,439 males). Post-matching analysis demonstrated adequate balance in covariates with standardized mean differences in age and categorical distributions below 0.1% (or below 2% between matched sex groups). Demographic characteristics of AD and matched control patients are shown in Table 1.

**Table 1.**
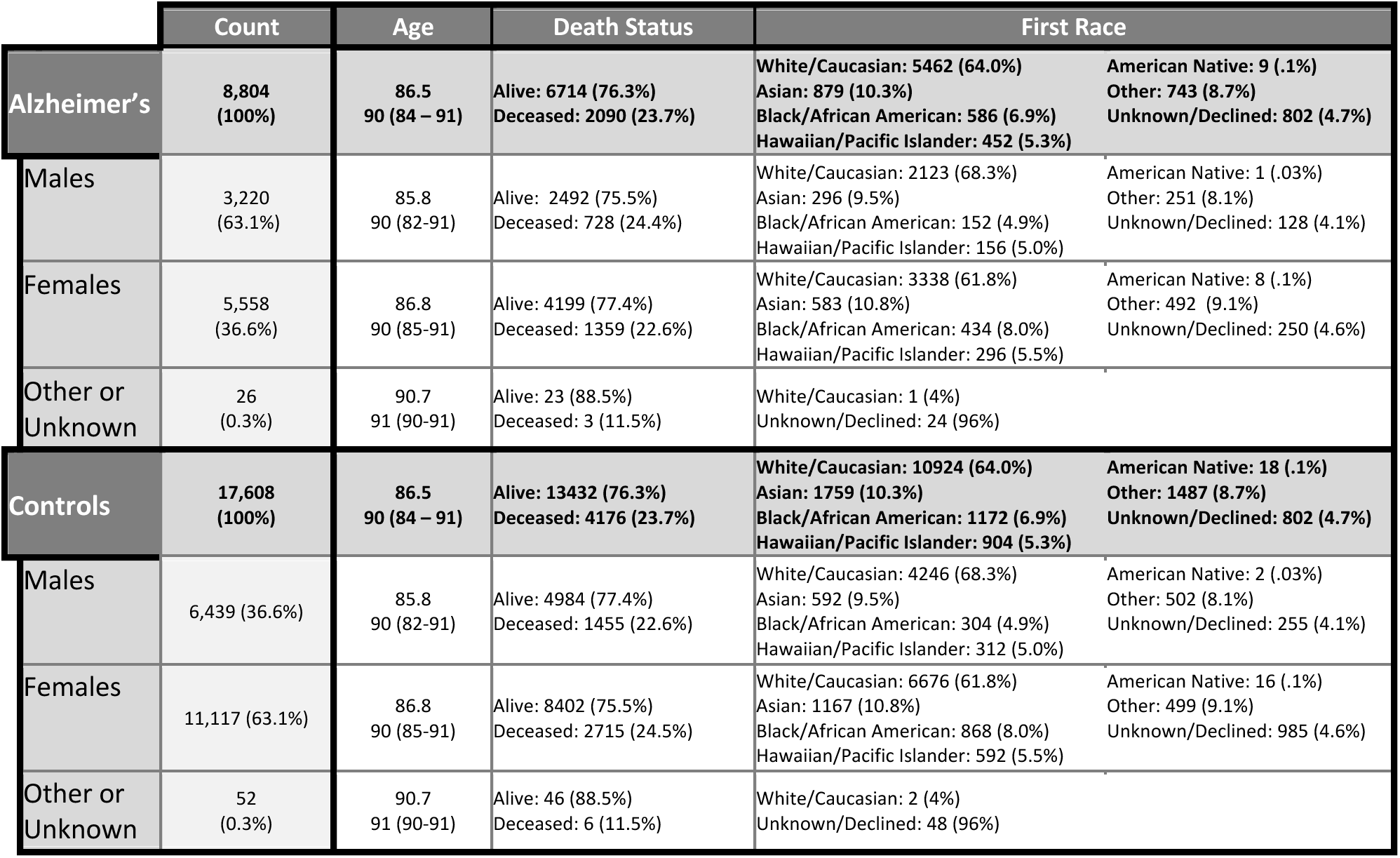
Patient Demographics. Distribution of sex, estimated age, death status, and first race among Alzheimer’s and control cohorts. Patients are matched at a 1:2 Alzheimer to control ratio with the demographics shown in the table. Estimated age shows mean and median (25%ile - 75%ile).

|  | Count | Age | Death Status | First Race |  |
| --- | --- | --- | --- | --- | --- |
| **Alzheimer's** | **8,804**<br>**(100%)** | **86.5**<br>**90 (84 – 91)** | **Alive: 6714 (76.3%)**<br>**Deceased: 2090 (23.7%)** | **White/Caucasian: 5462 (64.0%)**<br>**Asian: 879 (10.3%)**<br>**Black/African American: 586 (6.9%)**<br>**Hawaiian/Pacific Islander: 452 (5.3%)** | **American Native: 9 (.1%)**<br>**Other: 743 (8.7%)**<br>**Unknown/Declined: 802 (4.7%)** |
| Males | 3,220<br>(63.1%) | 85.8<br>90 (82-91) | Alive: 2492 (75.5%)<br>Deceased: 728 (24.4%) | White/Caucasian: 2123 (68.3%)<br>Asian: 296 (9.5%)<br>Black/African American: 152 (4.9%)<br>Hawaiian/Pacific Islander: 156 (5.0%) | American Native: 1 (.03%)<br>Other: 251 (8.1%)<br>Unknown/Declined: 128 (4.1%) |
| Females | 5,558<br>(36.6%) | 86.8<br>90 (85-91) | Alive: 4199 (77.4%)<br>Deceased: 1359 (22.6%) | White/Caucasian: 3338 (61.8%)<br>Asian: 583 (10.8%)<br>Black/African American: 434 (8.0%)<br>Hawaiian/Pacific Islander: 296 (5.5%) | American Native: 8 (.1%)<br>Other: 492 (9.1%)<br>Unknown/Declined: 250 (4.6%) |
| Other or Unknown | 26<br>(0.3%) | 90.7<br>91 (90-91) | Alive: 23 (88.5%)<br>Deceased: 3 (11.5%) | White/Caucasian: 1 (4%)<br>Unknown/Declined: 24 (96%) |  |
| **Controls** | **17,608**<br>**(100%)** | **86.5**<br>**90 (84 – 91)** | **Alive: 13432 (76.3%)**<br>**Deceased: 4176 (23.7%)** | **White/Caucasian: 10924 (64.0%)**<br>**Asian: 1759 (10.3%)**<br>**Black/African American: 1172 (6.9%)**<br>**Hawaiian/Pacific Islander: 904 (5.3%)** | **American Native: 18 (.1%)**<br>**Other: 1487 (8.7%)**<br>**Unknown/Declined: 802 (4.7%)** |
| Males | 6,439 (36.6%) | 85.8<br>90 (82-91) | Alive: 4984 (77.4%)<br>Deceased: 1455 (22.6%) | White/Caucasian: 4246 (68.3%)<br>Asian: 592 (9.5%)<br>Black/African American: 304 (4.9%)<br>Hawaiian/Pacific Islander: 312 (5.0%) | American Native: 2 (.03%)<br>Other: 502 (8.1%)<br>Unknown/Declined: 255 (4.1%) |
| Females | 11,117 (63.1%) | 86.8<br>90 (85-91) | Alive: 8402 (75.5%)<br>Deceased: 2715 (24.5%) | White/Caucasian: 6676 (61.8%)<br>Asian: 1167 (10.8%)<br>Black/African American: 868 (8.0%)<br>Hawaiian/Pacific Islander: 592 (5.5%) | American Native: 16 (.1%)<br>Other: 499 (9.1%)<br>Unknown/Declined: 985 (4.6%) |
| Other or Unknown | 52<br>(0.3%) | 90.7<br>91 (90-91) | Alive: 46 (88.5%)<br>Deceased: 6 (11.5%) | White/Caucasian: 2 (4%)<br>Unknown/Declined: 48 (96%) |  |

Low dimensional UMAP visualizations of non-AD diagnoses (47,439 features, ICD10 codes) show that distributions for AD and control patients are significantly different among the first two UMAP components (p-value < 1e-5, Figure 2A), with a progressive separation between groups. Sex and death status show significant differences along the first component, while age is significantly correlated among both components (Figure 2B, Supplementary Figure 1).

**Figure 1.**
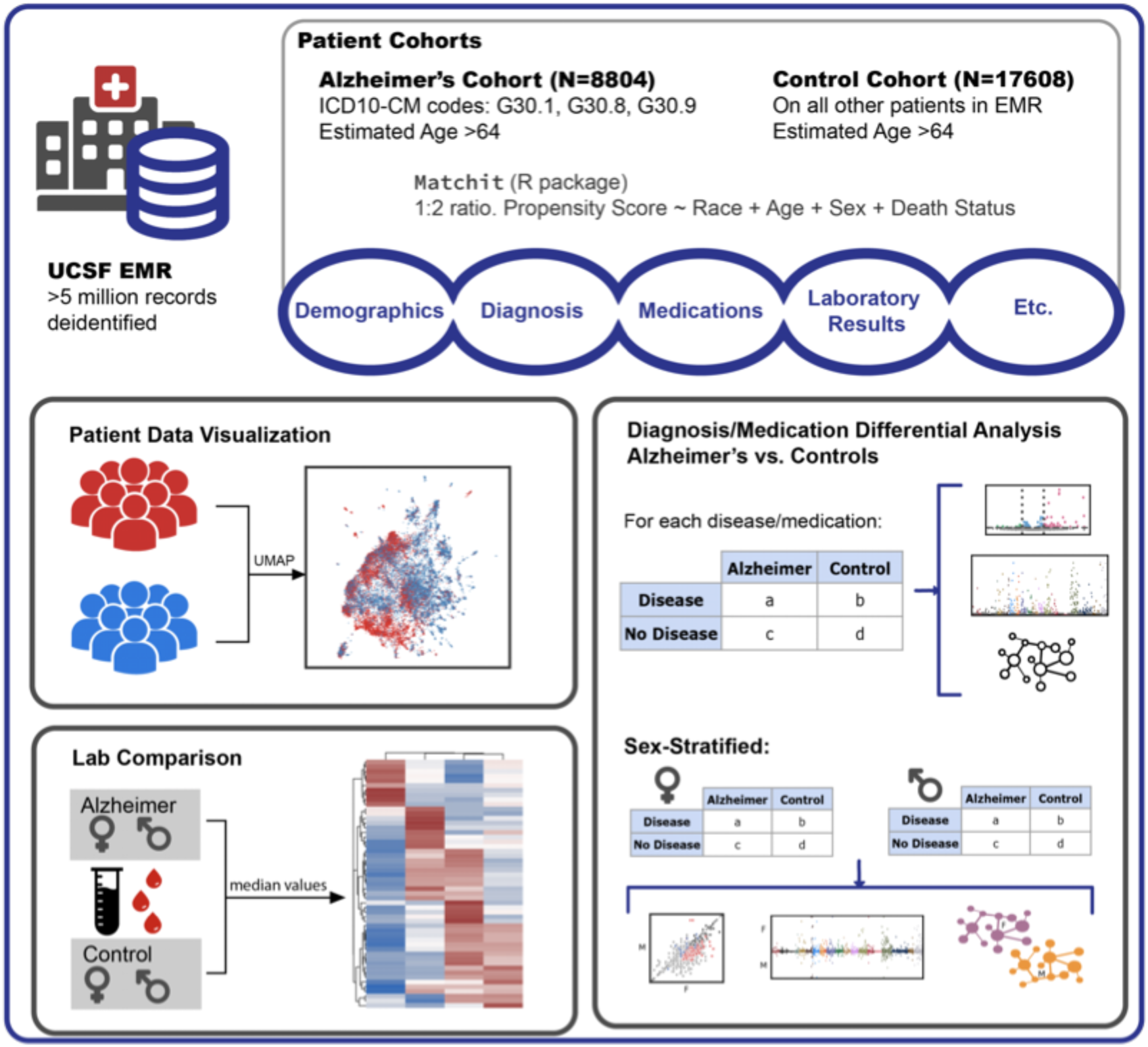
Workflow visualization. Visualization of patient cohort identification from the UCSF EMR and methods for deep phenotyping and enrichment analysis.

**Figure 2:**
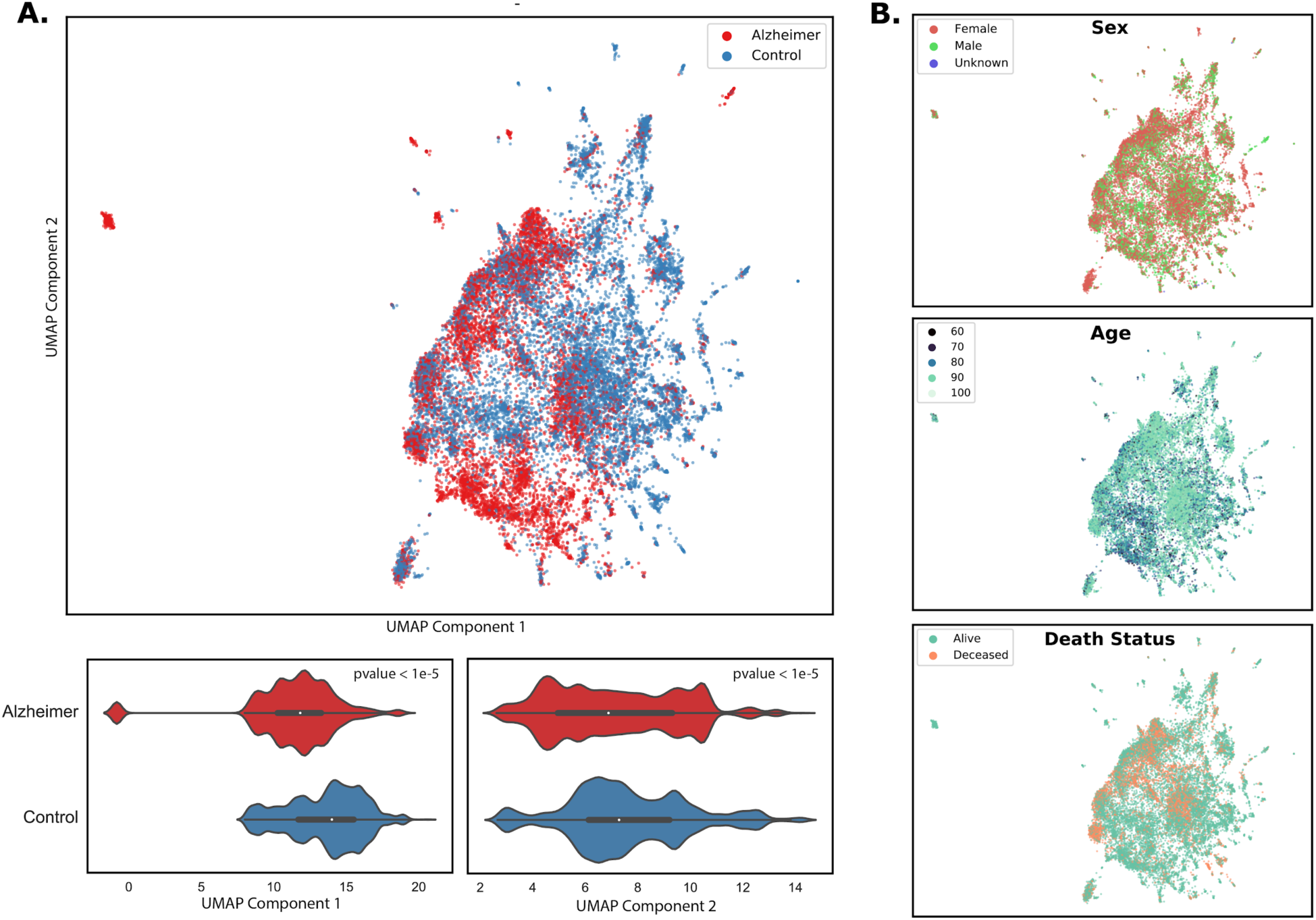
UMAPs using comorbidities as features provides a topographical view of the distribution of patients. Left: UMAP of all patients (AD and controls), with each dot representing a patient, colored by AD status. Bottom violin plots show distribution of AD and control patients along the UMAP principal components, and the distributions are compared with a Mann Whitney U Test. Right: UMAP of all patients colored by Sex, Age, and Death Status. Each dot represents a patient.

### Case Control Enrichment Analysis of Comorbidities

#### AD vs Controls

Among each diagnostic hierarchical level (Level 2 categories, Level 3 categories, and full diagnostic names), networks were constructed with each node representing a diagnostic category or diagnosis shared by >1% of patients in a group, and each edge representing a pair of diagnostic categories or diagnoses shared by >1% in a group. AD disease networks contain significantly more nodes and edges compared with control networks (full network metrics in Supplementary Table 3). When comparing node-level network metrics between groups, AD and control networks are significantly different (p-value < 0.01) when compared on average shortest path length, closeness centrality, neighborhood connectivity, and stress centrality, indicating a higher degree of connectivity among AD networks across all levels (Level 3 metrics in Figure 3C). When thresholding level 3 diagnostic categories by >5% of patients, the AD disease network has 243 diagnosis pairs compared to one pair in controls (Figure 3A).

**Figure 3:**
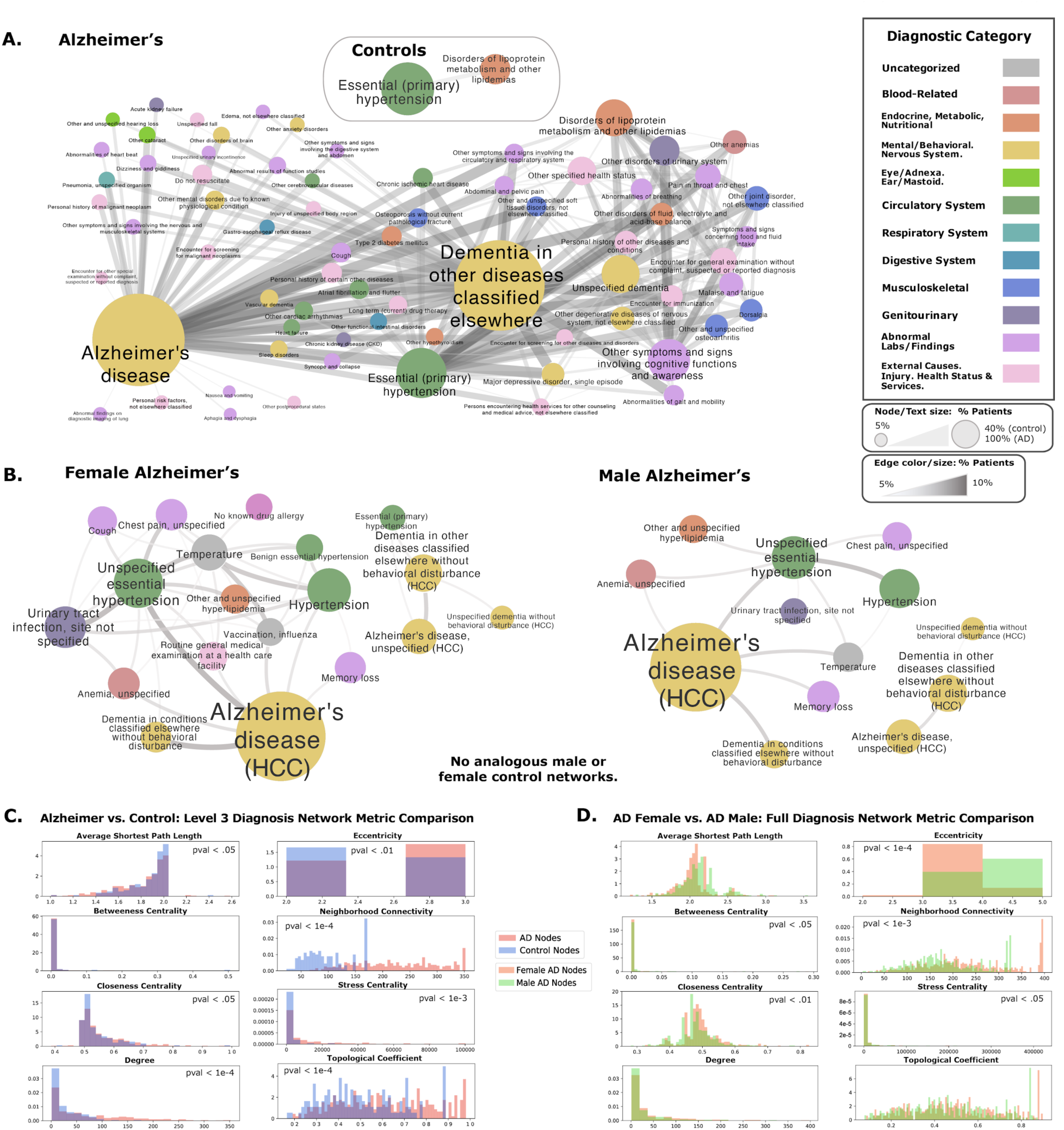
Comorbidity Networks Show Greater Co-Diagnosis in AD vs. Controls, and in Female AD vs Male AD patients. **A.** Network for level 3 diagnostic categories in AD vs. control patients. Nodes and edges represent >5% of diagnosis or diagnosis pairs shared in each cohort, respectively. **B.** Left: Female network of full diagnosis names. Each node and edge represent diagnosis or diagnosis pairs shared by >5% of AD females. No analogous comorbidity network was generated from control females. Right: Analogous network of diagnosis names for males. No network was produced on control males. For each network, the node size, text size, edge size, and edge color represent the number of patients sharing a diagnosis or diagnosis pair. Node colors are based on ICD10 category **C.** Comparison of network metrics between AD and control Level 3 Diagnostic Category Networks. Statistical Tests are performed with Mann Whitney U Test. **D.** Comparison of network metrics between Male and Female Alzheimer’s Disease Full Diagnostic Name Networks. Statistical Tests are performed with Mann Whitney U Test.

Within Level 2 diagnostic categories, there are 166 significant diagnostic categories (Bonferroni-corrected p-value < 0.05) identified to be different between AD and controls, with 120 significant diagnostic categories enriched (OR > 2) uniquely in the AD group (Figure 4A). Within Level 3 diagnostic categories, there are 501 significant categories, with 391 and 4 categories enriched in AD and control groups, respectively (Supplementary Table 2). Within full diagnosis names, there are 1627 significant diagnoses, with 1491 and 7 diagnoses enriched uniquely in AD and control groups, respectively. Top diagnoses in AD include vascular dementia, hypertension, hyperlipidemia, urinary tract infection, syncope, hypothyroidism, and osteoporosis, while top diagnoses in controls include neoplasms of liver and brain (Figure 4A, Supplementary Table 4). Top ICD diagnostic blocks in AD include mental health and behavioral diseases, genitourinary diseases, endocrine and metabolic diseases, and circulatory system diseases (Figure 4B).

**Figure 4.**
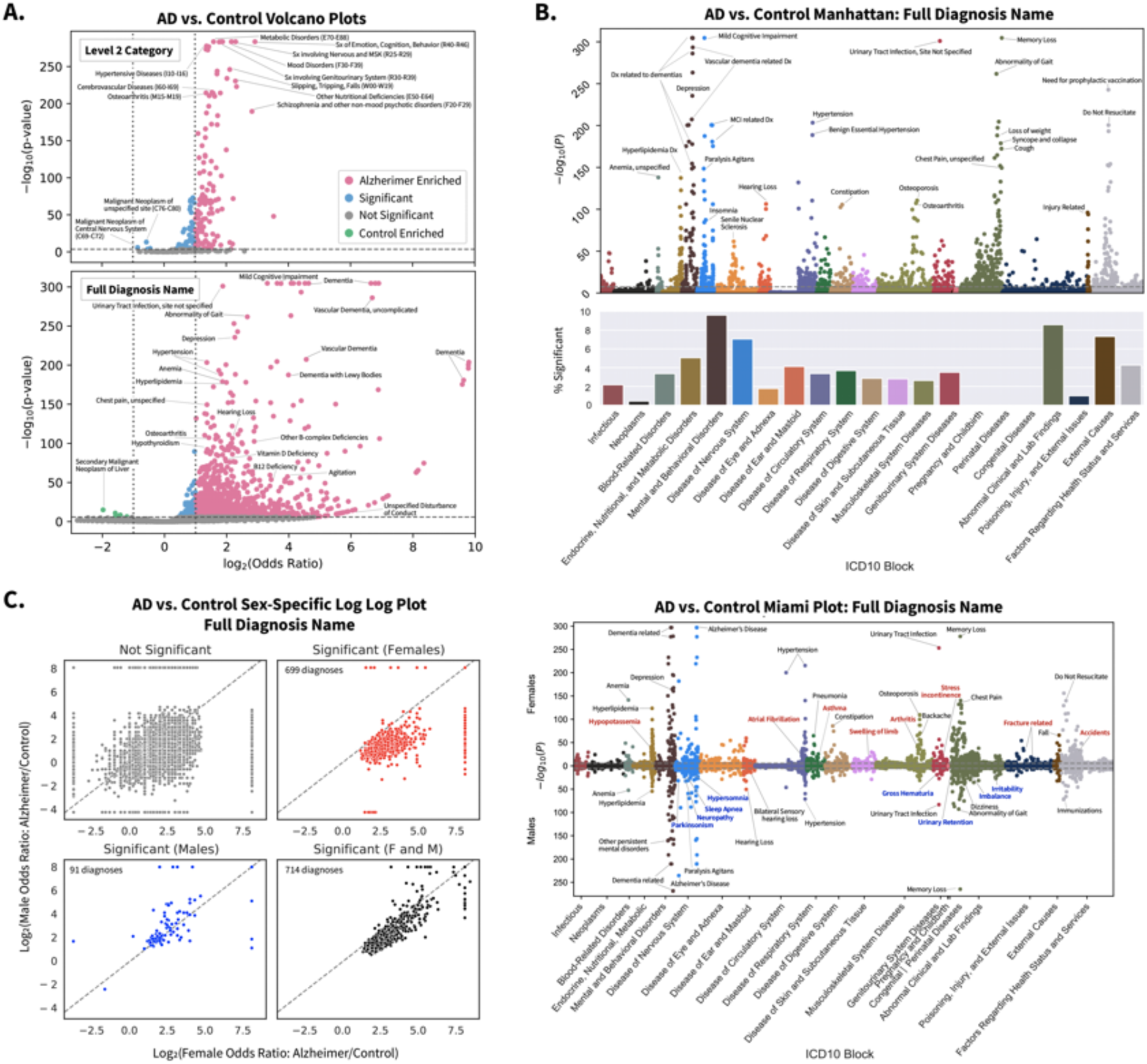
Comorbidity Enrichment Analysis identifies enriched diagnosis in AD vs. Controls as well as sex specific diagnostic enrichments. **A.** Volcano plot for level 2 categories (top) and full diagnosis names (bottom) compared between AD and control cohorts using Fisher Exact or Chi Squared test. P-value cutoff is Bonferroni corrected (p-val < 2e-8 and 1e-6) with odds ratio cutoff at 2 (or log odds ratio of 1) for AD enriched (pink) or 1/2 (or log odds ratio of -1) for control enriched (green), and remaining significant diagnoses in blue. **B.** Above, a Manhattan plot with full diagnosis names colored by ICD10 categories with Bonferroni p-value cutoff. Bottom, percentage of diagnosis in each ICD10 category that is significant. **C.** Full diagnosis names compared between AD and controls within each sex. The log of the odds ratio is plotted on the axis, and points are colored by significance (Bonferroni- corrected, p-val cutoff > 3e-6). **D.** Miami plot of the diagnosis names grouped by sex and ICD10 categories. Diagnosis names are colored by significance as female only (red), male only (blue), or significant in both sexes (black).

#### Sex-Stratified Case Control Enrichment Analysis of Comorbidities

When stratifying diagnoses by sex (see Methods), AD disease networks are significantly different on metrics of betweenness centrality and neighborhood connectivity in both males and females compared to their respective controls among all diagnostic levels. Networks were significantly different in stress centrality among all diagnostic levels when phenotyping AD males to AD females, but not when comparing control males to control females. Comparison of sex specific network for Diagnosis Name show greater closeness centrality, greater neighborhood connectivity, and lower eccentricity in female networks (Mann-Whitney U Test, p-value<0.01 all three metrics, Figure 3D, Supplementary Table 3). When thresholding full diagnosis names by >5% of patients within a sex group, female AD patients have 45 shared co-diagnosis pairs compared to 14 in male AD patients (Figure 3B, Supplementary Table 3), and no pairs were identified for either control sex group.

For both males and females, there are 136, 338, and 714 shared significant diagnostic categories or diagnoses for L2, L3, and full diagnosis names, respectively. In a sex-stratified analysis, there were 29, 164, and 699 female-only significant hits and 5, 18, and 91 male-only significant hits for L2, L3, and full diagnosis names (Figure 4C, Supplementary Table 4).

Compared to males among L2 diagnostic categories, females have a greater percent of significant diagnoses in blood-related disorders and congenital disorders and also have greater enrichment of pervasive and specific developmental disorders, musculoskeletal disorders (e.g. chondropathies, other osteopathies), injuries (e.g. injuries to the hip and thigh, injuries to the ankle and foot), infections with a predominantly sexual mode of transmission, and metabolic disorders (Supplementary Table 4).

Within full diagnosis names, unique significant diagnoses of female AD patients include asthma, atrial fibrillation, arthritis, fractures, and accidents while unique significant diagnosis of male AD patients include parkinsonism, sleep apnea, hypersomnia, neuropathy, irritability, and imbalance (Figure 4C, Figure 4D, Supplementary Table 4). Among diagnoses significant in both males and females, female AD patients have greater association in depression, hypertension, hyperlipidemia, urinary tract infections, upper respiratory infections, anemia, osteoporosis, and pneumonia, while male AD patients have greater importance in dementias with behavioral issues, hearing loss, and agitation (Supplementary Table 4).

#### Sensitivity Analysis Taking Encounters Into Account

We carried out a sensitivity analysis by thresholding AD patients on number of encounters ≥ 10 and records in EMR spanning > 1 year, resulting in 6,612 AD patients (2,382 males, 4,223 females). A second control cohort was identified analogously with additional matching on number and timespan of encounters, resulting in 13,224 control patients (4,674 males, 8,539 females). Demographics of this cohort are shown in Supplementary Table 1.

When thresholding and controlling on the number of encounters and total years recorded in EMR, we see 100, 222, and 561 significant L2, L3, and Full Diagnosis names respectively (Bonferroni-corrected p-value threshold of 0.05). After encounter controlling, there is an increase in odds ratio for chromosomal abnormalities and cerebrovascular disorders in AD patients (Supplementary Table 5). With sex-stratified enrichment analysis, encounter controlling increased enrichment of cerebrovascular disease in females, and increased significant enrichment of behavioral disorders, vision problems, and vascular dementia in males (Supplementary Table 5).

#### Rshiny App

An interactive visualization of Figure 3 and Figure 4 are made available in an RShiny app <u>vizad.org</u>.

### Case Control Medication Enrichment Analysis

In addition to comorbidities, we performed medication enrichment analysis in order to phenotype patients and investigate drugs enriched in controls. Medications found enriched (Bonferroni-corrected p-value < 0.05, OR >2 or < .5) in AD patients include current treatments like donepezil and memantine, but also vitamin B12, antidepressants (escitalopram, citalopram, sertraline, mirtazapine, trazodone), antipsychotics (quetiapine, risperidone, olanzapine), carbidopa/levodopa, vitamin D3, and melatonin. Medications found enriched in control patients include dexamethasone and those with lesser effect size (0.5 < OR < 1) including opioids and furosemide (Figure 5A).

**Figure 5:**
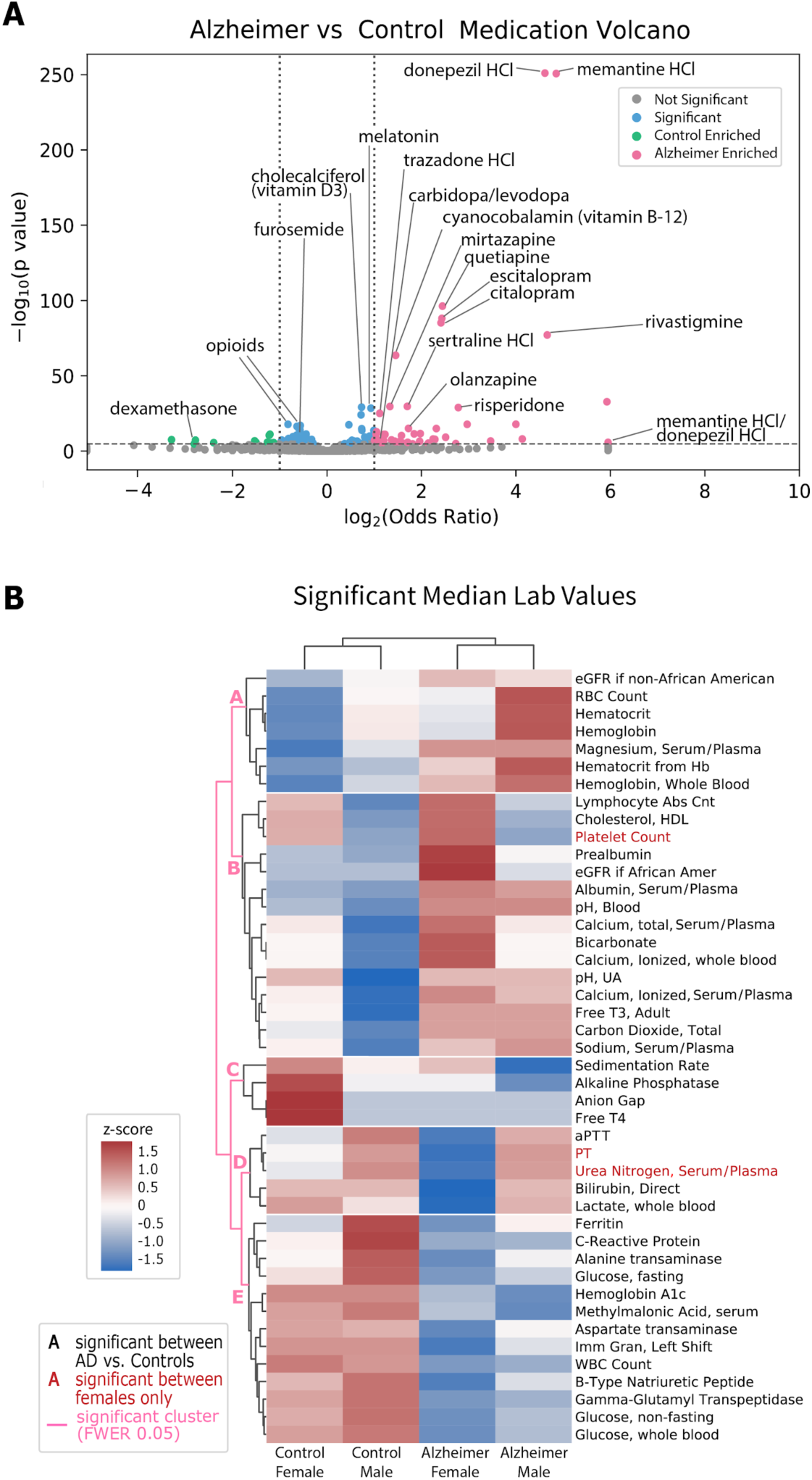
Medication and Lab Analysis shows Medication Enrichments and Median Lab Value Differences. **A.** Volcano plot for generic medication names compared between AD and controls using Fisher Exact or Chi Squared Test. P-value cutoff is Bonferoni corrected (p-val < 2e-5) with odds ratio cutoff at 2 for AD enriched (pink) or 1/2 for controlled enriched (green). Remaining significant diagnosis are in blue. **B.** Heatmap of lab values filtered on significance in case-control comparison across sex-specific groups. Labs are clustered with pink lines representing significant cluster breaks (FWER corrected p-value 0.05). Text color represents significant labs when compared among females only (red), or significant among AD vs. controls (black). Heatmap colors represent z-score of the average median value across the 4 groups.

### Lab Analysis

We also performed an unbiased analysis of laboratory test result differences between cases and controls in order to phenotype patient groups for diagnostic considerations and for identifying ‘opposite’ phenotypes among control groups. Median lab values were computed for every patient, and compared between groups with Mann Whitney U test (Bonferroni-corrected p-value threshold of 0.05). Among significantly different median lab values, AD patients have higher levels of hematocrit, serum calcium, RBC count, serum albumin, and C-reactive protein while controls have greater non-fasting glucose, white blood cell count, alanine transaminase, and aspartate transaminase (Figure 5C).

Average significant median lab values across sex-stratified groups (AD females, AD males, control females, control males) were clustered into 5 significant clusters (FWER corrected p-value 0.05 cutoff, Figure 5C). Clusters A and B represent groups where lab values are higher in AD compared to controls, but in Cluster A males have lab values higher than or equal to females within each group, and in Cluster B females have lab values higher or equal compared to males in each group. In Cluster A, AD Males have phenotypes of greater hemoglobin, hematocrit, and magnesium. In Cluster B, AD Females have phenotype of greater prealbumin, albumin, HDL Cholesterol, and blood pH, and platelet count is greater among Female AD vs Female controls only. Cluster B also shows control males with phenotype of lower calcium, bicarb, urine pH, free T3, carbon dioxide, and serum sodium.

In Cluster C, control females have lab values greater compared to all other groups, while cluster D contains labs where AD females have lower values compared to other groups. Cluster C shows control females with phenotype of greater sedimentation rate, alkaline phosphatase, anion gap, and free T4. Cluster D shows AD females with lower aPTT, Direct Bilirubin, and blood Lactate, and PT and BUN lower when compared between females only.

Lastly, cluster E contains lab values where AD labs are generally greater than control labs, with less sex variation. Cluster E shows control Males with phenotype of greater Ferritin, C-reactive protein, and alanine transaminase. Cluster E also generally shows control patients with phenotype of greater glucose, hemoglobin A1C, WBC Count, B-type natriuretic peptide, and methylmalonic acid.

## Discussion

In this work, we demonstrate the capability of utilizing data from EMRs in order to perform deep phenotyping of a complex and heterogeneous disease, Alzheimer’s Disease, and derive insights into associations with AD in a combined and sex-stratified analysis.

First, we performed low dimensional topographical embedding of patients using diagnoses as features in order to visualize patients spatially. We see that AD status is significantly correlated with the two UMAP components, suggesting that phenotypic representation of patients using diagnosis data can demonstrate separation of patients. The UMAP representation demonstrates a progressive spectrum between controls and AD, as well as representing variance and heterogeneity at individual patient resolution. Furthermore, with the UMAP representation, we can visualize topographically the distribution of age, sex, and other variables among patients.

We then generated comorbidity networks between AD and controls which provide a phenotypic representation of disease interactions among patient groups and a difference in connectivity between diseases in AD and controls. AD networks contain a greater number of edges and network metrics that point to higher rates of comorbid conditions among AD patients, particularly with stronger links of hypertension(HTN)-hyperlipidemia, HTN-urinary tract infection(UTI), and HTN-anemia. Indeed, other studies have found multimorbidities (such as neuropsychiatric and cardiovascular patterns) to increase risk for dementia^32^, and to contribute to AD pathological heterogenity^33, 34^ displaying the larger complexity and heterogeneous nature of AD.

With enrichment analysis, we apply an integrative, unbiased, big data approach to identify previously known associations and possible novel connections with AD. Some diagnoses found enriched in our analysis that have been previously identified as linked with AD include midlife hypertension^13, 35^, diabetes mellitus^15, 36^, anemia^37, 38^, vascular pathology^14, 39^, osteoporosis^40, 41^, and urinary tract infections^42^. Enrichment of hypertension and vascular risk factors supports many current hypotheses of potential vascular pathologies and inflammatory factors that may lead to AD^14, 43–45^ or ‘unmask’ the symptoms of AD by decreasing cognitive reserve by causing vascular brain disease. Enrichment of diabetes and dyslipidemia supports existing literature that found links with diabetes mellitus, with proposed hypotheses involving energy metabolism^46–48^, inflammation^49–51^, or related to integrity of blood brain barrier^52–54^. Enrichment of degenerative diseases of age, such as osteoporosis, osteoarthritis, urinary issues, and sensory issues may align with theories of AD as being a disease linked with frailty^55–57^. This analysis therefore provides an unbiased integrative way to identify multifactorial associations with AD. Our enrichment analysis also identified neoplasms as enriched in controls, especially cancer of brain and liver. This supports ideas that cancer and AD co-occur less frequently than the general population^58, 59^. Some theories propose that AD and cancer have similar mechanisms and molecular pathways, but are dysregulated in different directions^60, 61^.

Next, we generated sex-specific comorbidity networks to provide insight into sex differences in the complexity of the disease. Female AD networks contain more nodes with network metrics suggesting greater connectivity than female controls or male AD networks. This may support association with greater combined diagnoses and multimorbidity in female AD patients compared to males. These associations either support greater risk of dementia in females as a result of multiple diseases, or support the theory of greater cognitive and pathological resilience to AD in females due to taking on a greater burden of more comorbidities. Furthermore, sex-stratified networks show secondary interactions between comorbidities and AD, such as links of HTN-UTI and HTN-chest pain among female AD populations, but not in male AD patients. These findings give higher order comorbidity interactions associated with AD that have not been examined previously.

When performing enrichment analysis, we identify sex-specific enrichments that may be linked to AD that have not been previously explored in depth. Male AD patients show enrichment of neurological and sensory disorders (sleep disorder, parkinsonism, and irritability), and among diagnosis significant in both sexes, male AD patients have stronger associations with behavioral diagnoses, agitation, and hearing loss. Prior studies have found hearing loss to increase risk of dementia diagnosis^62, 63^ or cognitive decline^64, 65^ in men. The enrichment of behavioral and neurological disorders found in male AD patients may indicate lessened resilience or higher occurrence of co-pathology. Furthermore, this analysis found the psychiatric phenotype associated with AD to be related to behavioral phenotypes in males compared to females, which is consistent with prior studies^66, 67^.

Female AD patients have enrichment of unique significant diagnosis in musculoskeletal categories (arthritis, fractures), atrial fibrillation, and accidents, and among diagnoses significant in both sexes, female AD patients show stronger association with depression, hypertension, urinary tract infections, and osteoporosis. These diagnoses support vascular risk factors and pathology that may affect females more. Indeed, there is evidence supporting cardiovascular fitness to be protective or vascular risk factors to be harmful towards cognitive decline and dementia in women^35, 68–70^. Furthermore, these diagnoses suggest a phenotype for female AD patients along with other degenerative diseases of aging and frailty. In particular, the increase in musculoskeletal and bone disorders in female AD patients, as well as hypopotassemia and vitamin D deficiency, may point to a potential bone metabolism pathology or aberrant calcium metabolism in female AD patients. From a psychiatric standpoint, the female AD phenotype is more associated with depression compared to males as supported by studies that found depression associated with greater hippocampal volume loss in women^71^, and is more likely to be a manifestation of mild cognitive impairment or AD in females^72, 73^.

We performed sensitivity analysis by taking the number of encounters for each group into account. In general, we see a decrease in statistical significance in our enrichment analysis consistently across all diagnoses. This is likely due to decreased power from a lower sample size, and a bias towards selection of patients with more severe disease due to encounter thresholding. Overall, enriched diagnoses are relatively similar, with an increase in cerebrovascular disorders observed in AD, and particularly female AD patients. Neuroimaging studies have identified differences in AD phenotypes and brain networks depending on presence of cerebrovascular disease^74, 75^, which may support cerebrovascular events as an associated phenotype for a different or severe phenotype of AD.

Medication enrichments show expected associations with AD, as the top medication hits are current therapies used to modify symptoms of AD (e.g. memantine, donepezil), or are associated with diagnoses found in comorbidity analysis (e.g. antidepressants for depression). Medications enriched in controls provide a more interesting story, as it not only shows an ‘opposite AD’ phenotype, but control-enriched hits may provide a way to identify potential targets for further exploration of protective drug effects or drug repurposing. From our medication analysis, we see control enrichments of opioids, sedatives, dexamethasone, and furosemide. The negative association with opioids decreases support for prior studies that found associations between prescription opioid use and AD risk^76^, although control enrichment of opioids could possibly be due in part to decreased ability to communicate pain and decreased opioid prescriptions after AD^77^. Nevertheless, studies have implicated the role of opioid system dysregulation in tau hyperphosphorylation and AD^78^. Dexamethasone is a corticosteroid that has been suggested to help reduce inflammation in AD^79, 80^, although the data on efficacy is still uncertain and may depend upon the need for combination therapy^81^ or control of other factors that complicate the relationship between hormonal levels and the brain^82, 83^. Furosemide is a diuretic drug used to treat hypertension, and may confer a protective effect through the control of comorbid conditions that contribute to vascular risk factors. Furosemide also reduces the production of CSF by inhibiting carbonic anhydrase, which may impact CSF dynamics and help decrease the risk of AD^84^. Prior studies have shown possible protective effects from diuretic drugs and AD^85–87^, and one study identified furosemide as a potential probe molecule for reducing neuroinflammation^88^.

Characterizing patients by lab values provides another way to phenotype patient groups. These lab associations may provide a way to help identify AD risk early depending on the extent of similarity between a patient’s lab value to those of an AD subgroup. Greater calcium levels, especially in AD females, may support associations with osteoporosis and female sex modifying AD phenotype risk. Although one small observational study found calcium supplementation to increase risk of dementia in women with cerebrovascular disease^89^. Nevertheless, calcium dysregulation and homeostasis have been implicated in AD neuronal signaling pathology, and identified as a target for drug development^89, 90^. Control-enriched labs may also be related to gastrointestinal cancers or liver/pancreatic dysfunction, as we observe increased AST, ALT, and glucose levels in controls. This result is not consistent with a study observing greater glucose levels to increase dementia risk^91^, although one study did find low ALT^92^ to be associated with AD, and some publications implicate altered glucose metabolism^93, 94^ and liver dysfunction in AD pathology^92, 95, 96^. Furthermore, since our control cohort has been matched on age and death status, control patients may encompass a population with more morbid diseases. Lab clusters also demonstrate phenotypes specific to a sex group. A lower clotting time (aPTT, PT), bilirubin, and lactate and greater platelet count, prealbumin, and cholesterol levels in female AD patients may provide a multivariate way to identify potential AD phenotype in females. Prior studies have shown high thrombin^97, 98^, abnormalities of hemostasis^99, 100^, and abnormal platelet activation^101–,103^ in AD patients that may contribute to a pro-thrombotic state in AD^104^, leading to microinfarcts and cerebrovascular dysfunction^105, 106^, although sex specific associations have not been studied previously. Furthermore, control sex phenotype may demonstrate protective labs or biomarkers that decrease risk of AD. We see lower free T3 in control males, and greater free T4 in control females. Indeed, studies on AD populations have shown high TSH and low free T4 to be associated with the disease^107–109^, although sex specific associations have not been explored in depth.

Some limitations do exist with our study. First, AD is an insidious and heterogeneous disorder, and is frequently misdiagnosed even in specialized dementia centers. Our current study did not rely on biomarker positive cases of AD, and we did not exclude patients with other pathologies that can also impact brain health through different pathways, such as vascular dementia. Second, EMRs, while a rich data source, is a very sparse dataset with a lot of missing data. Nevertheless, the number of patients represented in the EMR is exceptionally large and provides robust opportunities for deriving meaningful insights or hypotheses. Third, our analysis only identifies associations with AD and does not take temporal factors into consideration, therefore causal relationships cannot be concluded. This will be the main focus of future work, as the temporal association can categorize an association as a risk/protective factor (if early in age), a diagnostic clue (if during AD diagnosis), or as a manifestation of AD progression or severity (if after AD diagnosis). Nevertheless, given AD is an insidious disorder, there can be brain perturbations a decade or more before a diagnosis is determined and documented in clinical records.

Overall, our analyses leverage an extensive clinical dataset to (1) phenotype and represent AD and (2) perform enrichment analysis to identify known or suggested novel associations with AD, as well eliciting sex-specific differences. We are therefore able to provide an integrative, unbiased, big data approach to identify associations with AD and controls, and provide phenotypic representations of an otherwise complex disease. With this approach, we can generate many new hypotheses to better motivate future work to understand AD complexity, diagnostic strategies and therapeutic interventions. Future work will include temporal analysis in order to identify longitudinal relationships, and perform predictive modeling for AD risk, diagnosis, or progression. Furthermore, more extensive analysis of medication and lab values, especially among opposite phenotypes in controls, may lead to better strategies for prevention or treatment of AD.

## Conclusion

We performed an integrative analysis of electronic medical record data in order to perform deep phenotyping of AD patients and identify sex-specific associations to extract insight into clinical characteristics of the disease. Low dimensional embeddings and network representation of diagnosis allow visualization of AD clinical heterogeneity and complexity. Enrichment analysis between cases and controls identifies known or new clinical associations in an unbiased manner, such as AD associations with hypertension, hyperlipidemia, and urinary tract infection and negative associations with neoplasms, opioid prescriptions, and diuretic drugs. Performing enrichment analysis in a sex-stratified manner allows us to identify novel sex-specific associations that can be further investigated to understand how sex modifies AD risk and phenotype. We found association of depression, osteoporosis, cardiovascular risk factors, and musculoskeletal disorders in female AD patients, and associations of neurological, behavioral, and sensory disorders in male AD patients. With lab analysis, we found greater calcium and lower ALT in AD, as well as abnormal hemostasis labs in female AD. Our analysis therefore provides an integrative and unbiased approach to identify associations with AD and provide phenotypic representations of a complex and heterogeneous disease. This analysis can be used to generate hypothesis that can be further investigated to help researchers and clinicians better understand AD complexity and sex-specific differences.

## Methods

We perform deep phenotyping and association analysis of AD and control patients. First, AD and control cohorts are identified from the UCSF electronic medical records and topographically visualized via a low dimensional projection of comorbidities. Comorbidity networks are created, and association and enrichment analysis was performed on all diagnoses, medications, and lab values. These analyses were further performed in a sex-stratified manner to identify sex specific associations. An overview of the workflow is shown in Figure 1.

### Patient Identification

Patient cohorts were identified from over five million patients in the UCSF EMR database. Due to the de-identification process, dates are shifted by at most a year (with relative dates preserved) and all patients over 90 years of age are represented as 90 years old. AD patients were identified by inclusion criteria of estimated age >64 years, and ICD10 codes G30.1, G30.8, or G30.9. We identified a control group within patients >64 years old without AD diagnosis using propensity score (PS) matching (*matchit* R package^110^) on sex, estimated age, race, and death status at a 1:2 AD:control ratio using a nearest neighbors method. Propensity score was estimated by the logistic regression.

### Dimensionality Reduction Patient Visualization

All identified patients are represented using one-hot encoding of diagnosis. All diagnoses with Alzheimer’s in the name were removed (list in Supplementary Table 2). Patients were then visualized in a lower dimension using Uniform Manifold Approximation and Projection^111^ (UMAP) with umap-learn package from Python. Correlations between variables and UMAP coordinates were analyzed using Mann-Whitney U-Test for categorical variables, and Pearson’s Correlation Coefficient for continuous variables.

### Case Control Enrichment Analysis of Comorbidities

To evaluate comorbidities, all diagnoses recorded from patient cohorts were identified with the earliest entry of every diagnosis. Comparisons were made at different ICD10 hierarchical levels, specifically Level 2 categories (L2 name, e.g. G30-G32: Other degenerative diseases of the nervous system), Level 3 categories (L3 name, e.g. G30: Alzheimer’s Disease), or full diagnosis names (e.g. G30.9 Alzheimer’s Disease, unspecified) and grouped by ICD10 blocks (e.g. G00- G99: Diseases of the Nervous System. More information on ICD10 codes can be found here: https://www.cms.gov/Medicare/Coding/ICD10/ICD-10Resources).

Diagnosis networks were created based upon a diagnostic category or diagnosis shared by >1% of a group (node) or pair of categories/diagnoses shared by >1% of a group (edge). Network metrics were computed using Cytoscape app Network Analyzer^15^. Metrics were then compared between AD and control networks using Mann-Whitney U-test, with and without singleton nodes removed. Nodes and edges were thresholded by 5% of patients for visualization purposes.

Enrichment analysis of diagnosis was compared between AD and control cohorts. For each diagnosis, the proportions of patients in each group were compared using Fisher Exact (if <5 patients in a category) or Chi-Squared test. Significant diagnoses were determined by a Bonferroni-corrected threshold of p-value < 0.05, and directionality determined with Odds Ratio (OR). With inspiration from genetic and molecular approaches, the results are visualized using Manhattan plots by categorizing diagnoses in ICD10 blocks.

### Sex-Stratified Case Control Enrichment Analysis of Comorbidities

Diagnostic networks were created for each sex, with diagnosis categories or diagnoses shared by >1% of a group (node), and diagnostic category/diagnosis pair shared by >1% of a group (edge). Network metrics were then computed using Cytoscape Network Analyzer app, and compared between sex stratified cases and controls, and between males and females for both AD and controls separately with a Mann-Whitney U-test. Nodes and edges were thresholded by 5% of patients for visualization.

Sex specific enrichment analysis of diagnoses between AD and control cohorts were compared with a subset of equal numbers of male and female patients. For each diagnosis, the proportions of patients in each group was compared using the Fisher Exact (if <5 patients in a category) or Chi Squared test. Significance was determined by applying a threshold of 0.05 for Bonferroni-corrected p-values. Log-log plots were generated from odds ratios between Female and Male case and controls, and Miami plots created by categorizing diagnoses in ICD10 blocks.

### Sensitivity Analysis Taking Encounters Into Account

Sensitivity analysis of diagnosis enrichment analysis was performed with a subgroup of AD patients and a second control cohort to take into account variability in the number of visits for each patient. AD cohorts were subgrouped by identifying patients with over 10 encounters in the EMR and records spanning over a year. The encounter-controlled control cohort was identified by additionally matching on the number of encounters and years between the first and last record in the EMR. Diagnostic enrichment analysis was carried out as described above for general comorbidities and sex-specific analysis.

### Case Control Enrichment Analysis of Medications

All medications ordered for AD and control patients were extracted and grouped based upon the generic medication name, with route and dosage information removed. The proportions of AD and control patients prescribed each medication were compared using Fisher Exact (if <5 patients in a category) or Chi Squared tests. Significantly enriched medications were identified by a Bonferroni-corrected threshold of p-value 0.05, and directionality was determined with an Odds Ratio (OR <0.5 or >2).

### Case Control Comparisons of Lab Values

For laboratory values, median values for all numerical lab test results for each patient were identified. Lab tests missing data among 95% or more patients were removed. Lab value distributions were compared using Mann Whitney U-test across three comparisons (AD vs. controls, Female AD vs. Female controls, and Male AD vs. Male controls) in order to identify significantly different lab values.

For clustering analysis, significant lab tests above a threshold of 0.05 for Bonferroni-corrected p-value were isolated, and mean values were then identified for each group (AD Females, AD Males, control Females, control Males) and normalized across groups as a Z-score. Clustering was then performed using the *sigclut2* R package^112^ to determine significance of each cluster break using permutations (Euclidean distance metric and average linkage).

### Data Visualization Using RShiny

An interactive visualization of comorbidity enrichments and networks between case and control groups and with sex stratification was implemented in an Rshiny^113^ app: <u>vizad.org</u>.

## Data Availability

A visualization of comorbidities and networks can be found at https://vizad.org. Comorbidity tables can also be found in the supplement.

## Acknowledgements

Primary support through Grant # NIA R01AG060393, R01AG057683 (A.T., T.O., C.W.S., M.S.). Additional support provided by NIA RF1AG068325 (D.B.D) and Medical Scientist Training Program T32GM007618 (A.T.). We’d like to acknowledge Zachary Cutts and Stella Belonwu for their suggestions and help.

## Author Contributions

A.T. and M.S. designed question, experiments, and analytic plan. B.Z. and Z.H. helped with data acquisition, cleaning, and interpretation. A.T., C.W.S., and B.O. helped with creation of Rshiny app. A.T., C.W.S., W.M., and D.D. interpreted results. A.T. wrote the manuscript with editing from all the authors. All the authors edited and reviewed the manuscript.

## Supplementary Figures and Tables

**Supplementary Table 1.** **Patient Demographics with Encounter Thresholds and Controlling.** Distribution of sex, estimated age, death status, and first race among Alzheimer’s and control cohorts. These cohorts are threshold-ed on more than 10 encounters, and over a year representation in the EMR. Patients are matched at a 1:2 Alzheimer to control ratio with the demographics shown in the table. Estimated age shows mean and median (25%ile - 75%ile).

|  |  | Count | Age | Death Status | First Race |
| --- | --- | --- | --- | --- | --- |
| **Alzheimer's** |  | **6,612**<br>**(100%)** | **86.4**<br>**90 (83-91)** | **Alive: 6714 (76.3%)**<br>**Deceased: 2090 (23.7%)** | **White/Caucasian: 5462 (64.0%)**<br>**Asian: 879 (10.3%)**<br>**Black/African American: 586 (6.9%)**<br>**Hawaiian/Pacific Islander: 452 (5.3%)**<br>**American Native: 9 (.1%)**<br>**Other: 743 (8.7%)**<br>**Unknown/Declined: 802 (4.7%)** |
|  | Males | 2,382<br>(36.0%) | 85.7<br>90 (84-91) | Alive: 1778 (74.6%)<br>Deceased: 604 (25.4%) | White/Caucasian: 1570 (66.8%)<br>Asian: 254 (10.8%)<br>Black/African American: 128 (5.4%)<br>Hawaiian/Pacific Islander: 117 (5.0%)<br>American Native: 1 (.04%)<br>Other: 209 (8.9%)<br>Unknown/Declined: 72 (3.1%) |
|  | Females | 4,223<br>(63.9%) | 86.8<br>90 (82-91) | Alive: 3084 (73%)<br>Deceased: 1139 (27%) | White/Caucasian: 2525 (60.5%)<br>Asian: 497 (11.9%)<br>Black/African American: 393 (9.4%)<br>Hawaiian/Pacific Islander: 217 (5.2%)<br>American Native: 8 (0.2%)<br>Other: 404 (9.7%)<br>Unknown/Declined: 130 (3.1%) |
|  | Other or Unknown | 7<br>(0.1%) | 90.7<br>91 (90.5-91) | Alive: 7 (100%) | White/Caucasian: 6 (85.7%)<br>Unknown/Declined: 1 (14.3%) |
| **Controls** |  | **13,224**<br>**(100%)** | **86.2**<br>**90 (83 – 91)** | **Alive: 13432 (76.3%)**<br>**Deceased: 4176 (23.7%)** | **White/Caucasian: 10924 (64.0%)**<br>**Asian: 1759 (10.3%)**<br>**Black/African American: 1172 (6.9%)**<br>**Hawaiian/Pacific Islander: 904 (5.3%)**<br>**American Native: 18 (.1%)**<br>**Other: 1487 (8.7%)**<br>**Unknown/Declined: 802 (4.7%)** |
|  | Males | 4,674<br>(35.3%) | 85.8<br>90 (82-91) | Alive: 3248 (69.5%)<br>Deceased: 1426 (30.5%) | White/Caucasian: 3076 (66.8%)<br>Asian: 490 (10.7%)<br>Black/African American: 277 (6.0%)<br>Hawaiian/Pacific Islander: 222 (4.8%)<br>American Native: 5 (.1%)<br>Other: 384 (8.3%)<br>Unknown/Declined: 247 (3.2%) |
|  | Females | 8,539<br>(64.6%) | 86.5<br>90 (84-91) | Alive: 6253 (73.2%)<br>Deceased: 2286 (26.8%) | White/Caucasian: 5225 (61.9%)<br>Asian: 1024 (12.1%)<br>Black/African American: 768 (9.1%)<br>Hawaiian/Pacific Islander: 387 (4.6%)<br>American Native: 11 (.1%)<br>Other: 783 (9.3%)<br>Unknown/Declined: 246 (2.9%) |
|  | Other or Unknown | 11<br>(0.1%) | 90.2<br>90 (90-91) | Alive: 10 (90.9%)<br>Deceased: 1 (9.1%) | White/Caucasian: 1 (11.1%)<br>Other: 3 (33.3%)<br>Unknown/Declined: 5 (55.6%) |

**Supplementary Table 2. UMAP Exclusion Terms** Csv file of diagnosis excluded in UMAP embedding. These terms contain the word ‘Alzheimer’.

**Supplementary Figure 1.**
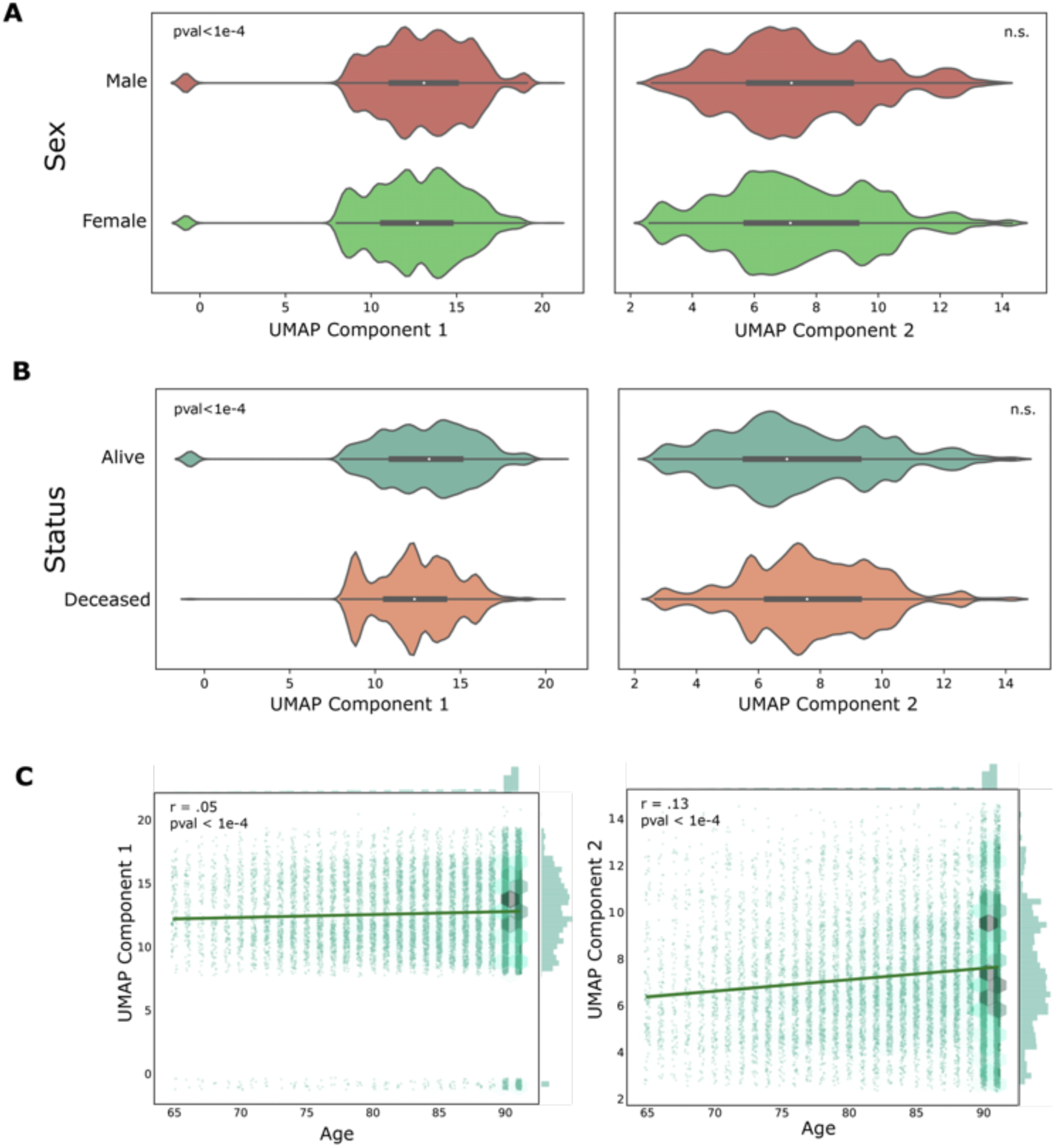
Demographic correlation across UMAP principal components. **A.** The two graphs show the distribution of sex across the two UMAP principal components. Distributions are compared with a Mann Whitney U Test. **B.** The two graphs show distribution of death status across two UMAP principal components. Distributions are compared with Mann Whitney U Test. **C.** The two plots show the distribution of the estimated age across the two UMAP principal components. Darker hexagons show higher relative density of patients, with marginal distribution shown on the sides. There is greater density around 90 years old due to the de-identification process. A regression line is plotted, and a Pearson’s R correlation test is performed.

**Supplementary Table 3. All Diagnosis Network Metrics**

An attached excel sheet with 2 tabs.

Tab 1: Diagnosis Networks are created with nodes representing a diagnostic category or diagnosis shared among >1% of patients in a group, and edges representing >1% of co-diagnosis in a group. These networks are then analyzed for overall network metrics shown in the table.

Tab 2: Network metrics are computed for nodes in each network, and the distribution of metrics are compared between networks. Comparisons are performed with and without the removal of singletons (single nodes with no neighbors). A Mann Whitney U test is performed to compare the distribution of each network metric, with colors based upon p-value cutoff.

**Supplementary Table 4. Thresholded Full Tables of Diagnosis Enrichment Analysis.** 1 excel sheet will be attached with 3 levels of diagnostic names and sex-specific analysis (6 tabs). Lists include diagnosis enriched between AD and controls, and sex-specific enrichments. Diagnosis are threshold-ed to represent > 10 patients, with p-values and odds-ratios. The data can be visualized and explored in the Rshiny app: <u>vizad.org</u>.

**Supplementary Table 5. Encounter Controlled Diagnosis Enrichment Analysis.** 1 excel sheets will be attached for the 3 levels of diagnostic names on encounter controlled control cohorts (described in Methods) and sex-specific analysis (6 tabs0. Lists include diagnosis enriched between AD and controls, and sex-specific enrichments. Diagnoses are threshold-ed to represent > 10 patients, with p-values and odds-ratios. The data can be visualized and explored in the Rshiny app: <u>vizad.org</u>.

