## Supplementary Table 2 for "Deep clinical phenotyping of Alzheimer’s Disease Patients Leveraging Electronic Medical Records Data Identifies Sex-Specific Clinical Associations"

AD (Alzheimer's disease) (HCC)
Alzheimer disease (HCC)
Alzheimer disease type 3 (HCC)
Alzheimer's dementia (HCC)
Alzheimer's dementia with behavioral disturbance (HCC)
"Alzheimer's dementia with behavioral disturbance, unspecified timing of dementia onset (HCC)"
Alzheimer's dementia without behavioral disturbance (HCC)
"Alzheimer's dementia without behavioral disturbance, unspecified timing of dementia onset (HCC)"
"Alzheimer's dementia, late onset (HCC)"
"Alzheimer's dementia, late onset, with behavioral disturbance (HCC)"
Alzheimer's disease (HCC)
Alzheimer's disease of other onset
Alzheimer's disease of other onset with behavioral disturbance (HCC)
Alzheimer's disease of other onset without behavioral disturbance (HCC)
Alzheimer's disease with delirium (HCC)
Alzheimer's disease with early onset (CODE) (HCC)
Alzheimer's disease with early onset (HCC)
Alzheimer's disease with late onset (CODE) (HCC)
Alzheimer's disease with late onset (HCC)
Alzheimer's disease with presenile onset (HCC)
"Alzheimer's disease, early onset (HCC)"
"Alzheimer's disease, familial (HCC)"
"Alzheimer's disease, focal onset (HCC)"
"Alzheimer's disease, unspecified (CODE) (HCC)"
"Alzheimer's disease, unspecified (HCC)"
Alzheimer's type dementia (HCC)
Alzheimer's type dementia with late onset with behavioral disturbance (HCC)
Alzheimer's type dementia with late onset without behavioral disturbance (HCC)
Alzheimers disease (HCC)
DAT (dementia Alzheimer type)
DAT (dementia of Alzheimer type) (HCC)
Dementia due to Alzheimer's disease (HCC)
Dementia in Alzheimer's disease (HCC)
Dementia in Alzheimer's disease with delusions (HCC)
Dementia in Alzheimer's disease with depression (HCC)
Dementia in Alzheimer's disease with early onset (HCC)
Dementia in Alzheimer's disease with early onset with behavioral disturbance (HCC)
Dementia in Alzheimer's disease with early onset without behavioral disturbance (HCC)
"Dementia in Alzheimer's disease with early onset, with behavioral disturbance"
"Dementia in Alzheimer's disease with early onset, without behavioral disturbance"
Dementia in Alzheimer's disease with late onset
Dementia of Alzheimer's type with behavioral disturbance (HCC)
"Dementia of Alzheimer's type, with early onset, with depressed mood (HCC)"
Dementia of the Alzheimer's type (HCC)
Dementia of the Alzheimer's type with early onset with behavioral disturbance (HCC)
Dementia of the Alzheimer's type with late onset without behavioral disturbance (HCC)
Dementia of the Alzheimer's type without behavioral disturbance (HCC)
"Dementia of the Alzheimer's type, with late onset, uncomplicated (HCC)"
"Dementia of the Alzheimer's type, with late onset, with delirium (HCC)"
"Dementia of the Alzheimer's type, with late onset, with delusions (HCC)"
"Dementia of the Alzheimer's type, with late onset, with depressed mood (HCC)"
"Dementia of the Alzheimer's type, with late onset, with depressive mood (HCC)"
"Dementia, Alzheimer's, with behavior disturbance (HCC)"
Early onset Alzheimer disease
Early onset Alzheimer's dementia without behavioral disturbance (HCC)
Early onset Alzheimer's disease with behavioral disturbance (HCC)
Family history of Alzheimer's disease
Focal Alzheimer's disease (HCC) 'Late onset Alzheimer disease (HCC)
Late onset Alzheimer's disease with behavioral disturbance (HCC)
Late onset Alzheimer's disease without behavioral disturbance (HCC)
Major neurocognitive disorder due to Alzheimer's disease (HCC)
"Major neurocognitive disorder due to Alzheimer's disease, possible (HCC)"
"Major neurocognitive disorder due to Alzheimer's disease, probable, with behavioral disturbance (HCC)"
"Major neurocognitive disorder due to Alzheimer's disease, probable, without behavioral disturbance (HCC)"
"Major neurocognitive disorder due to Alzheimer's disease, with behavioral disturbance (HCC)"
Major neurocognitive disorder due to possible Alzheimer's disease (HCC)
"Major neurocognitive disorder, due to Alzheimer's disease, with behavioral disturbance, mild (HCC)"
"Major neurocognitive disorder, due to Alzheimer's disease, without behavioral disturbance, mild (HCC)"
"Major neurocognitive disorder, due to Alzheimer's disease, without behavioral disturbance, moderate (HCC)"
"Major neurocognitive disorder, due to Alzheimer's disease, without behavioral disturbance, severe (HCC)"
Mild major neurocognitive disorder due to Alzheimer's disease with behavioral disturbance (HCC)
Mild major neurocognitive disorder due to Alzheimer's disease without behavioral disturbance (HCC)
Mild neurocognitive disorder due to Alzheimer's disease (HCC)
Mild possible major neurocognitive disorder due to Alzheimer's disease (HCC)
Mixed Alzheimer's and vascular dementia (HCC)
Mixed Alzheimer's and vascular dementia with behavior disturbances (HCC)
Moderate major neurocognitive disorder due to Alzheimer's disease without behavioral disturbance (HCC)
Moderate probable major neurocognitive disorder due to Alzheimer's disease with behavioral disturbance
Other Alzheimer's disease (HCC)
Possible major neurocognitive disorder due to Alzheimer's disease
Primary degenerative dementia of Alzheimer type (HCC)
"Primary degenerative dementia of the Alzheimer type, senile onset (HCC)"
"Primary degenerative dementia of the Alzheimer type, senile onset, uncomplicated (HCC)"
"Primary degenerative dementia of the Alzheimer type, senile onset, with depression (HCC)"
Probable major neurocognitive disorder due to Alzheimer's disease with behavioral disturbance
Probable major neurocognitive disorder due to Alzheimer's disease without behavioral disturbance
Progressive aphasia in Alzheimer's disease (HCC)
SDAT (senile dementia of Alzheimer's type) (HCC)
Senile dementia of Alzheimer's type (HCC)
Sporadic Alzheimer's disease (HCC)'
